# Whole genome sequencing detects minimal clustering among *Escherichia coli* Sequence Type 131 *H30* isolates collected from U.S. children’s hospitals

**DOI:** 10.1101/19010140

**Authors:** Arianna Miles-Jay, Scott J. Weissmann, Amanda L. Adler, Janet G. Baseman, Danielle M. Zerr

**Author notes:** Address correspondence to: Dr. Arianna Miles-Jay, University of Michigan Medical School, 1150 W. Medical Center Dr., Medical Science Research Building I, Room 1511, Ann Arbor, MI 48109. A. Miles-Jay’s affiliation has changed since the completion of this work. Her current affiliation is Department of Microbiology and Immunology, University of Michigan, Ann Arbor, Michigan, USA.

## Abstract

*Escherichia coli* sequence type 131 *H30* has garnered global attention as a dominant antimicrobial-resistant lineage of extraintestinal pathogenic *E. coli*, but its transmission dynamics remain undefined. We applied whole genome sequencing to identify putative transmission clusters among clinical isolates of *H30* from children across the U.S. Of 126 isolates, 17 were involved in 8 putative transmission clusters; 4 clusters involved isolates with some evidence of healthcare-associated epidemiologic linkages. Geographic clustering analyses showed weak geographic clustering. These findings are consistent with a framework where within-hospital transmission is not a main contributor to the propagation of *H30* in a pediatric setting.

## Background

Extraintestinal pathogenic *Escherichia coli* (ExPEC) cause a wide range of non-intestinal illnesses, ranging from uncomplicated urinary tract infection to potentially fatal bacteremia [1]. Unlike intestinal pathogenic *E. coli*, which is commonly associated with outbreaks, ExPEC infections are historically considered sporadic, and tracking ExPEC transmission has not been a clinical or public health priority. However, the widespread dissemination of antimicrobial-resistant lineages such as sequence type (ST) 131-*H30* (also known as ST131 Clade C), a dominant multidrug-resistant (MDR) ExPEC lineage in both adults and children, has brought new interest to understanding the transmission dynamics of these common pathogens [2,3]. In particular, the relative importance of healthcare vs. community transmission to the propagation of MDR ExPEC lineages remains poorly defined, though recent work indicates community exposures may be more important in pediatric patients.[4]

The transmission dynamics of ST131-*H30* (hereafter, *H30*) are challenging to study. Like other ExPEC lineages, *H30* is known to asymptomatically colonize the gut for extended periods of time prior to—or potentially without ever—transitioning to extraintestinal infection [5]. These instances of long-term intestinal colonization likely result in numerous “silent” transmission events [6]. Whole genome sequencing (WGS) and phylogenetic methods can shed light on pathogen transmission dynamics even in the presence of silent transmission events. Here, we used WGS to identify putative transmission clusters among passively collected clinical *H30* isolates from four children’s hospitals across the U.S. We also quantified genomic evidence of geographic clustering to characterize the spatiotemporal dynamics of *H30* among children.

## Methods

### Strain collection and whole genome sequencing

All isolates and clinical data came from a previously described multicenter case-control study. Briefly, between September 1, 2009 and September 30, 2013, four freestanding children’s hospitals—referred to here as “West,” “Midwest 1,” “Midwest 2,” and “East”— collected *E. coli* isolates during the course of standard clinical care from individuals <22 years old. All extended-spectrum cephalosporin-resistant and a subset of extended-spectrum cephalosporin-sensitive isolates were collected [7]. The Institutional Review Board at each hospital approved the study protocol. *H30* isolates were identified using the *fumC/fimH* genotyping scheme [8]; only the first *H30* isolate per individual was included.

All *H30* isolates underwent WGS on the Illumina NextSeq platform. Sequencing reads were quality filtered, trimmed, mapped to a high-quality *H30* reference genome (EC958), and single nucleotide variants (SNVs) were called and filtered.[9] Filtered SNVs were used to construct a pairwise SNV distance matrix using snp-dists, and a maximum-likelihood phylogenetic tree, using IQ-tree [10,11]. See Supplementary Methods for more details. Sequence data generated are available from the NCBI Sequence Read Archive under BioProject PRJNA578285 (see Supplementary Table 2 for study sample metadata).

### Identification and characterization of putative clusters

Pairwise SNV distances within and between all combinations of collection sites were visualized, and the minimum SNV distance between two isolates from discordant collection sites was used to define a threshold for identification of putative transmission clusters. This approach was selected in an effort to capture direct and indirect transmission events that are epidemiologically relevant, i.e. would warrant further investigation should the clusters have been identified in real-time. Given the substantial geographic distance between the collection sites in this study, we expect no epidemiologically relevant transmission events that span two distinct collection sites. The transcluster package in R (version 3.5.1, R Core Team, 2018) was used to estimate counts of uncaptured transmission events separating isolates in each putative cluster while incorporating the sampling dates and estimated evolutionary rate and transmission rate of *H30* [12]. See Supplementary Methods for more details.

### Genomic evaluation of geographic clustering

To examine the spatiotemporal dynamics of *H30*, we quantified genomic evidence of geographic clustering using two approaches. The primary approach was a previously described SNV-distance based approach where a ratio of the median SNV distance within collection sites over the median SNV distance between collection sites (SNV_within_ / SNV_between_) is calculated, with a ratio closer to zero indicating more evidence of clustering by collection site [13]. Statistical significance of the pairwise-SNV distance-based clustering was assessed using permutation-based 95% interval estimates with 1000 permutations; a SNV distance ratio below the lower bound of the 95% interval estimate indicated evidence of geographic clustering. Secondarily, we applied a previously described phylogenetically informed approach [14]. Briefly, the number of isolates in well-supported phylogenetic clades of size 2 or greater that were homogeneous for collection site (“pure” clusters) was tallied. Statistical significance of these counts was again assessed using permutation-based 95% interval estimates with 1000 permutations; counts higher than the upper bound of the permuted 95% interval estimate were considered evidence of clustering. Both approaches were first executed on the full sample set and then on four temporally segregated sample sets in approximate one-year increments to explore whether geographic signal interacted with the temporal variability of sampling.

Clustering by “geographically close” sites compared to “geographically distant” sites was also examined. In SNV-distance based analyses, pairs of isolates that spanned Midwest 1 and Midwest 2 were classified as geographically close, while all other discordant site pairs were classified as geographically distant. In phylogenetically informed analyses, the two sites in the Midwest region of the U.S. were collapsed into one “Midwest” category. The same methods described above were applied to the full data set and to the temporally segregated sample sets.

## Results

One hundred thirty *H30* isolates were identified out of 1,347 *E. coli* screened over the four-year study period. Three of the 130 *H30* isolates were determined to be non-*H30* after *in silico* analysis and one isolate was identified as a subsequent isolate from an individual already in the study, leaving 126 *H30* isolates in the remainder of the analyses. After quality filtering and recombination masking, 3,433 variable sites were identified and included in the whole-genome-based SNV alignment.

There were 7,875 different pairwise comparisons made, with the pairwise SNV distance ranging from 0 to 165 SNVs. The minimum SNV distance between isolates from discordant collection sites was 14 SNVs, and pairs of isolates separated by less than or equal to 14 SNVs were considered to be members of a putative transmission cluster (Figure 1A). Using this threshold, eight putative clusters were identified involving seventeen isolates, seven clusters containing two isolates and one cluster containing three isolates (Figure 2A, Supplementary Figure 1). The putative cluster with three isolates (Cluster 1) consisted of one pair separated by 15 SNVs, which was just beyond the selected cutoff, but because the other two pairs within the cluster were separated by <14 SNVs, all three isolates were included in further analyses. Clusters contained a mix of community- and healthcare-associated isolates (Supplementary Figure 2).

**Figure 1:**
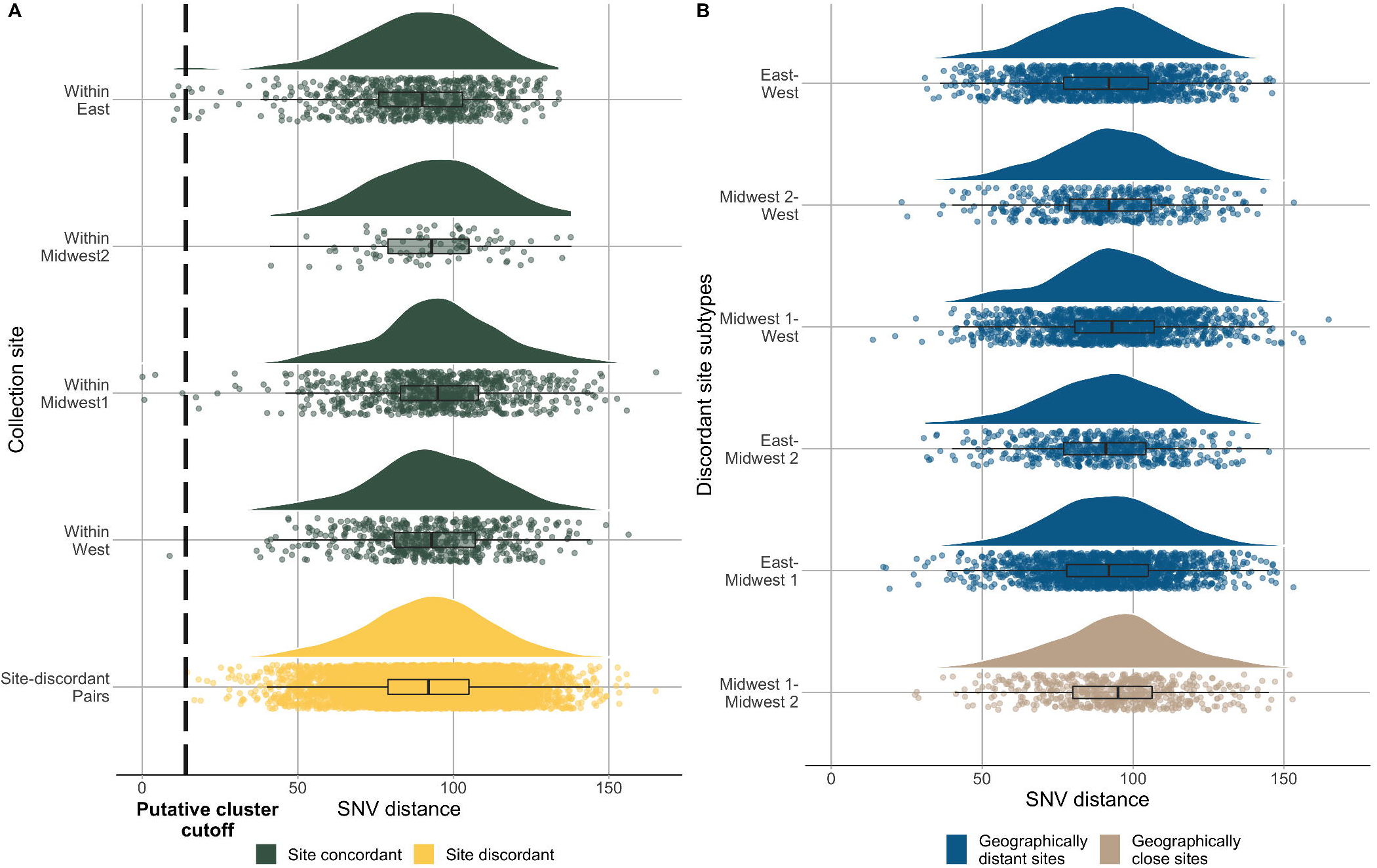
Distribution of pairwise single nucleotide variant (SNV) differences between *H30* clinical isolates from four children’s hospitals in the U.S. A) Within single collection sites compared to between discordant collection sites. Dashed line indicates selected threshold for identifying putative transmission clusters; and B) Between geographically distant discordant collection sites vs. between geographically close discordant collection sites.

**Figure 2:**
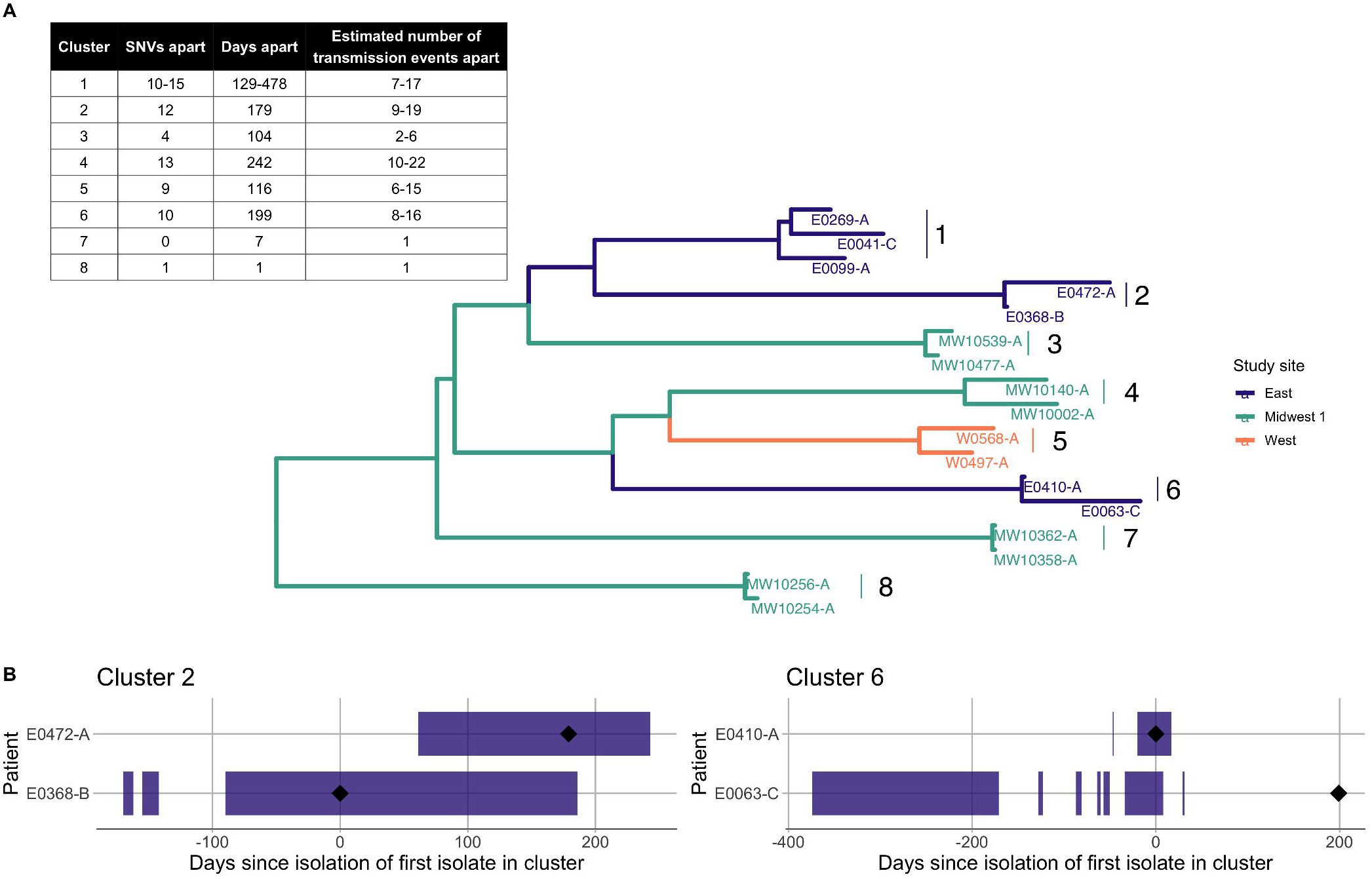
A) Phylogeny of eight identified putative transmission clusters of *H30* identified from four children’s hospitals in the U.S. colored by study site, with the number of days separating their collection; the number of single nucleotide variants (SNVs) separating them after quality filtering; and a range of estimated number of transmission events separating them as calculated by the R package transcluster. The range of estimated transmission events reflect a range of reasonable values for substitution and transmission rates, but do not account for potential intra-host evolution or diversity. B) Temporal depiction of the overlapping hospitalizations of individuals in Cluster 2 and Cluster 6. The black diamond indicates the date of isolate collection and the purple bars represent time hospitalized. Time is measured in days since the isolation of the first isolate in each cluster.

Out of the eight identified putative clusters, documented epidemiologic data associated with four clusters (Clusters 2,6,7,8) was consistent with possible nosocomial acquisition. Clusters 2 and 6 involved individuals with documented overlapping dates of hospitalization (Figure 2B). The genomic evidence for direct transmission within these clusters is less clear: they were separated by 12 and 10 SNVs, respectively, and the transcluster method estimated that there were between 8 and 19 transmission events separating the isolates in these clusters (Figure 2A). Additionally, only one of the two isolates in Cluster 2 was phenotypically resistant to trimethoprim-sulfamethoxazole (Supplementary Figure 2). However, the within-cluster difference in isolation dates was 179 and 199 days, so long-term colonization and within-host evolution may have inflated the estimated number of transmission events and resulted in a loss of a resistance determinant. Clusters 7 and 8 consisted of isolates that differed by 0-1 SNVs after quality filtering and were collected between 1 and 7 days of one another, with the transcluster method estimating direct transmission (Figure 2A). While there was no documentation of overlapping hospitalizations within Cluster 7 or 8, both individuals within Cluster 7 had surgical site infections associated with neurological procedures, while both individuals within Cluster 8 were paraplegic. These connections are consistent with a plausible epidemiological link in inpatient or outpatient care, although conclusively establishing such a link is outside the scope of these data.

Genomic clustering analyses demonstrated minimal evidence of geographic clustering by collection site. Visual inspection of geographic sites on the phylogenetic tree did not show remarkable evidence of geographic clusters (Supplementary Figure 1). The median SNV distance within pairs across concordant sites was not significantly different from the median SNV distance within pairs across discordant sites. (SNV_within_ / SNV_between_ = 1.01, 95% interval estimate 0.99-1.01 Figure 1A). Similarly, the median SNV distance within pairs across the most geographically proximate discordant sites (Midwest 1 and Midwest 2) was not significantly lower than the median SNV distance within pairs across more geographically distant site pairs (SNV_within_ / SNV_between_ = 1.03, 95% interval estimate 0.99-1.02, Figure 1B). The phylogenetic-informed approach demonstrated weak clustering in the data by study site and no clustering by Midwest sites vs. other sites (Supplementary Figure 3). Results for temporally segregated sample sets were similar, with most measures not supporting evidence of clustering (Supplementary Table 1 and Supplementary Figures 4 and 5).

## Discussion

We applied WGS to clinical isolates collected from four freestanding U.S. children’s hospitals over four years to identify putative transmission clusters and investigate the spatiotemporal dynamics of *E. coli* ST131-*H30*. We identified eight putative transmission clusters of *H30* involving seventeen isolates, including two clusters with documented overlapping hospitalizations and two clusters with other plausible healthcare-associated epidemiologic links. Genomic spatiotemporal analyses demonstrated little evidence of geographic clustering of *H30* more broadly.

To our knowledge, there are no available data about transmission of *H30* between children within healthcare settings. Our observation of limited plausible within-hospital transmission is not surprising, given the infrequent documentation of MDR-ExPEC transmission within healthcare, generally [15]. These findings are consistent with a framework where within-hospital transmission is not a dominant contributor to the propagation of *H30* in a pediatric setting. However, the identification of some plausible nosocomial transmission highlights the utility of WGS of isolates collected during the course of standard clinical care to uncover silent transmission events.

There are also no data, to our knowledge, describing the spatiotemporal dynamics of *H30* within the U.S. using geographically diverse isolates. Our observation of limited geographic clustering was unexpected; we anticipated a stronger genomic signature associated with sustained local circulation at the various geographic sites. These findings may reflect the rapid and recent dissemination of *H30* at the time of this data collection. Whether these patterns remain the same today, almost two decades after *H30* is believed to have disseminated rapidly and globally, is worthy of further study [2].

The results of this study should be interpreted in the context of multiple limitations. First, the available epidemiologic data were limited — including a lack of detail about the location of specific wards during overlapping hospitalizations — and, as such, all observations of plausible transmission should be interpreted cautiously. Second, as highlighted elsewhere [12], the selection of a SNV threshold as a method of defining putative transmission clusters has limitations. However, the multicenter design in this study provided the opportunity to apply a conservative threshold where even indirect transmission was epidemiologically unlikely, which we believe to be a reasonable approach given the limited transmission data available about *H30*. Finally, the local epidemiology of *H30* may have changed since the collection of these isolates. This study also had several strengths, including a multicenter design; a large collection of *H30* isolates from an understudied pediatric population; and the use of WGS and phylogenetically informed approaches to investigate both transmission-based and geographic clustering.

As antimicrobial resistance rates among ExPEC rise, there is new urgency to improve our understanding of the transmission dynamics of these common pathogens. Taken together, our findings of minimal evidence of transmission clusters or broader geographic clustering are consistent with the prevailing conceptualization of *H30* as a globally and recently disseminated epidemic strain that is often community-associated [2]. Future studies should consider focusing on community-based exposures when investigating the transmission dynamics of *H30*.

## Data Availability

Sequence data generated are available from the NCBI Sequence Read Archive under BioProject PRJNA578285

## Acknowledgements

The authors would like to thank Carey-Ann Burnham, Alexis Elward, Jason Newland, Rangaraj Selvarangan, Kaede Sullivan, Theoklis Zaoutis, and Xuan Qin for provision of bacterial isolates and associated clinical data; Brad Cookson for helpful advice; Jeff Myers and Huxley Smart for assistance with molecular typing of isolates; the Northwest Genomics Center for execution of whole genome sequencing; and members of the Snitkin Lab at the University of Michigan for their critical review of the manuscript.

